# Functional dysconnectivity of visual and somatomotor networks yields a simple and robust biomarker for psychosis

**DOI:** 10.1101/2024.06.14.24308836

**Authors:** Brian P. Keane, Yonatan Abrham, Michael W. Cole, Brent A. Johnson, Boyang Hu, Carrisa V. Cocuzza

**Affiliations:** Departments of Psychiatry and Neuroscience, University of Rochester Medical Center, 430 Elmwood Ave, Rochester, NY 14642, USA; Center for Visual Science, University of Rochester, 601 Elmwood Ave, P.O. Box 319, Rochester, NY 14642, USA; Department of Brain & Cognitive Science, University of Rochester, 358 Meliora Hall P.O. Box 270268, Rochester, NY 14627-0268, USA; Center for Molecular and Behavioral Neuroscience, Rutgers, The State University of New Jersey, 197 University Ave, 07102, USA; Department of Biostatistics, University of Rochester Medical Center, 601 Elmwood Ave, Rochester, NY, USA; Department of Psychology, Yale University, 100 College St, New Haven, CT 06510, USA

## Abstract

People with psychosis exhibit thalamo-cortical hyperconnectivity and cortico-cortical hypoconnectivity with sensory networks, however, it remains unclear if this applies to all sensory networks, whether it arises from other illness factors, or whether such differences could form the basis of a viable biomarker. To address the foregoing, we harnessed data from the Human Connectome Early Psychosis Project and computed resting-state functional connectivity (RSFC) matrices for 54 healthy controls and 105 psychosis patients. Primary visual, secondary visual (“visual2”), auditory, and somatomotor networks were defined via a recent brain network partition. RSFC was determined for 718 regions via regularized partial correlation. Psychosis patients— both affective and non-affective—exhibited cortico-cortical hypoconnectivity and thalamo-cortical hyperconnectivity in somatomotor and visual2 networks but not in auditory or primary visual networks. When we averaged and normalized the visual2 and somatomotor network connections, and subtracted the thalamo-cortical and cortico-cortical connectivity values, a robust psychosis biomarker emerged (p=2e-10, Hedges’ g=1.05). This “somato-visual” biomarker was present in antipsychotic-naive patients and did not depend on confounds such as psychiatric comorbidities, substance/nicotine use, stress, anxiety, or demographics. It had moderate test-retest reliability (ICC=.61) and could be recovered in five-minute scans. The marker could discriminate groups in leave-one-site-out cross-validation (AUC=.79) and improve group classification upon being added to a well-known neurocognition task. Finally, it could differentiate later-stage psychosis patients from healthy or ADHD controls in two independent data sets. These results introduce a simple and robust RSFC biomarker that can distinguish psychosis patients from controls by the early illness stages.

## Introduction

Psychiatry needs robust, generalizable biomarkers of psychosis. Such biomarkers could help clarify illness pathophysiology, predict illness onset, or stratify patients into clinically meaningful subgroups [1]. Here, we consider the possibility that functional dysconnectivity of sensory networks might provide such a marker. Past fMRI work has shown that–in later-stage schizophrenia –cortical sensory areas are more weakly connected to one another (‘hypoconnectivity’) and more strongly connected to the thalamus (‘hyperconnectivity’) [2]. The hyperconnectivity result has been replicated [3–6] but the hypoconnectivity result has received comparatively less attention. Moreover, these studies estimated connectivity via Pearson correlation, which cannot distinguish direct and indirect connections [7]. In many of these studies, various confounds were not ruled out and the specificity of the effect to psychosis remained an open question. Finally, no attempt has been made to coalesce these findings into a single marker.

To be clinically useful, a neuroimaging biomarker should have a number of features. It should: 1) be large in magnitude; 2) be robust to potential confounds including motion, medication, and comorbidities; 3) emerge with multiple preprocessing strategies [8]; 4) generalize to unseen data [9]; 5) differentiate psychosis patients from a clinical control group; 6) be recoverable from a relatively brief scan session; 7) complement and improve upon other more standard methods of discriminating groups (e.g., neurocognition); 8) have good retest reliability; and, ideally, 9) be biologically plausible and easy to interpret [e.g., 10]. We sought to establish such a marker by leveraging data from the Human Connectome Early Psychosis project. We focused on early psychosis patients since this population lacks illness chronicity confounds (e.g., poor health and diet, prolonged medication exposure). We restricted the hypothesis space in a principled way by conducting our analyses on four atlas-defined sensory networks [11] (Fig. 1). Moreover, our results were computed at the network level so as to yield potentially larger and more generalizable group differences [11, 12]. Finally, we derived resting-state functional connectivity (RSFC) matrices via regularized partial correlation (graphical lasso), which may offer the best strategy for removing spurious and indirect connections [13].

**Fig. 1.**
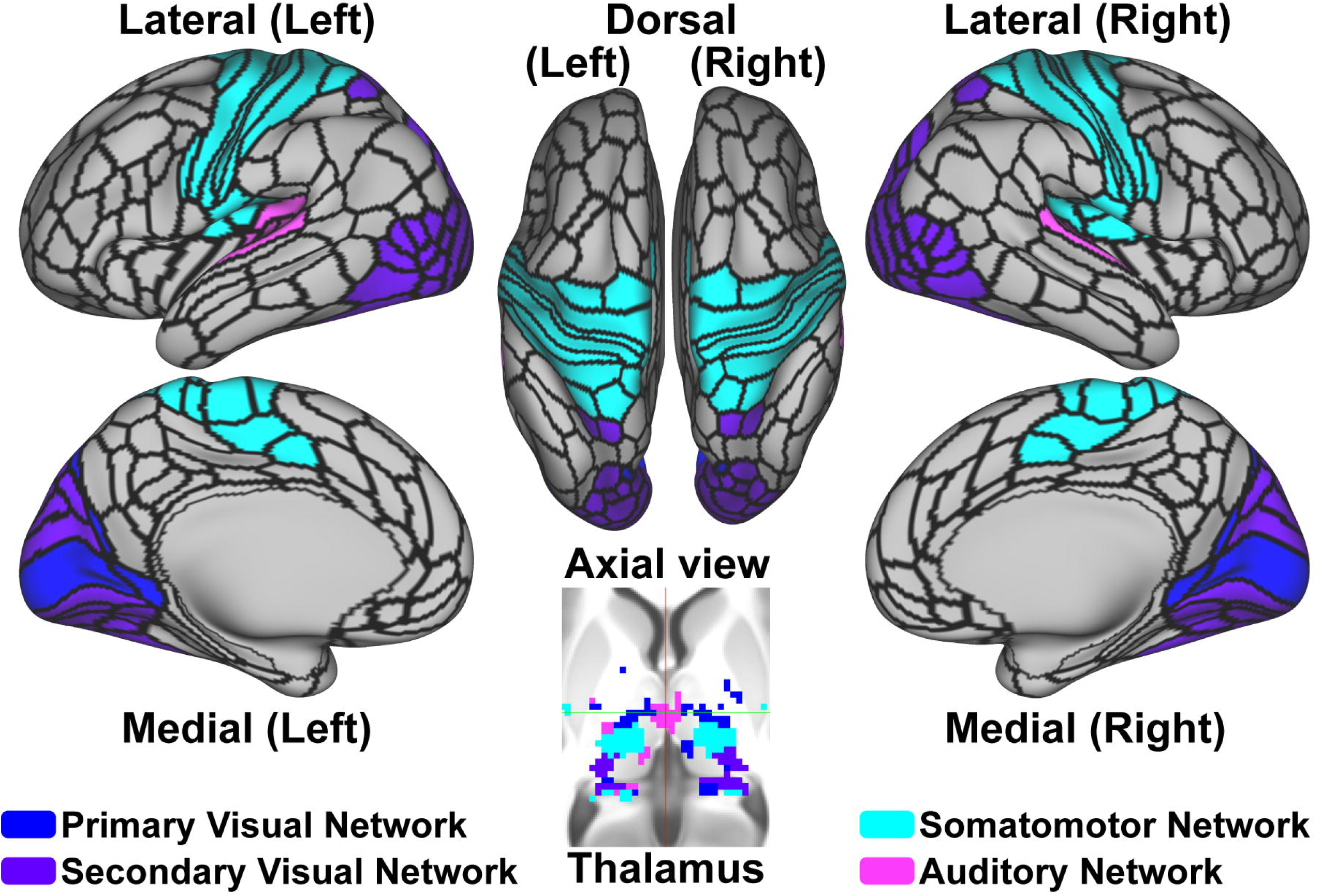
The four sensory networks of the brain network partition. The partition comprises four sensory networks, with the somatomotor network encompassing the somatosensory cortex. The thalamus contains parcels of each sensory network.

Below, we first generate RSFC matrices for each subject and identify the sensory networks that most obviously exhibit “dysconnectivity”, that is, abnormal cortico-cortical or thalamo-cortical connections [14]. Upon finding that the affective and non-affective groups were nearly the same on every RSFC measure and that they differed from controls on the somatomotor and secondary visual network, we combined these two groups and devised a novel “somato-visual” biomarker of psychosis. We show that this biomarker exemplifies most of the nine features enumerated above.

## Materials & Methods

### Participants

The HCP Early Psychosis Project (release 1.1) furnishes multimodal brain imaging data from healthy controls (n=54, 34 males, age_MEAN_=24.8), patients with nonaffective psychosis (n=81, 56 males, age_MEAN_=22.1), and patients with affective psychosis (n=24, 9 males, age_MEAN_=24.4; see Table S1 for further clinical and demographic details). This sample does not include 6 subjects (4 non-affective, 2 affective) who had a missing run, 6 subjects whose preprocessing failed or had otherwise low quality (1 control, 3 non-affective, 2 affective), and 3 non-affective patients with excessive in-scanner motion.

All patients had an illness onset within five years of testing. Diagnoses were based on the Diagnostic and Statistical Manual of Mental Disorders, Fifth Edition and were assessed via Structured Clinical Interview for DSM-5 (SCID) [15, 16]. Symptoms were assessed with the Positive and Negative Syndrome Scale [PANSS; 17] and a five-factor scoring system [18]. Medication dose on the day of the scan was recorded as chlorpromazine equivalents [19]. There were 22 patients who were antipsychotic-naive at the time of scanning.

### fMRI acquisition

T1w (MPRAGE) and T2-weighted (SPACE) scans were used for image preprocessing (slice thickness = 0.8 mm, 208 slices). Whole-brain multiband T2*-weighted echo-planar imaging (EPI) resting-state acquisitions were collected at four sites with 32- or 64-channel head coils. Data were not normalized according to testing site (or any other potential confound) at any stage of the analysis. There were four resting-state scans per subject (410 measurements; 5 minutes 28 seconds; TR=.8 s; voxel=2 mm^3^); these were acquired with eyes open and in alternating phase encoding directions (anterior-to-posterior, posterior-to-anterior; see Supplementary Methods).

### fMRI preprocessing and accounting for in-scanner motion

Imaging data were minimally preprocessed using fMRIPrep (Supplementary Methods). All subsequent preprocessing steps and analyses were conducted on CIFTI 91k grayordinate standard space using a parcellated time series (i.e., one BOLD time series for each parcel, averaged over grayordinates; see below for a description of the brain partition). We performed nuisance regression on the minimally preprocessed functional data using 24 motion parameters (6 motion parameter estimates, their derivatives, and the squares of each) and the 4 ventricle and 4 white matter parameters (parameter estimates, the derivatives, and the squares of each) [20]. Results were initially run without whole-brain global signal regression (GSR). When GSR was applied, we additionally included four more regressors (mean signal, its derivative, and the quadratic of each) [21]. Each run was also individually demeaned and detrended, adding 2 more regressors per run. To show robustness, a third preprocessing strategy– aCompCor–was also used; this incorporated the first five principal components of white matter and ventricles for the physiological regressors [22].

Additionally, we removed the first five frames of each run and applied motion scrubbing [23]. That is, whenever the framewise displacement for a particular frame exceeded 0.20 mm, we removed that frame, one prior frame, and two subsequent frames (Supplementary Methods). To reduce the effect of respiration on the framewise displacement measure, we applied a first-order Butterworth low pass (0.3 Hz) filter to the framewise displacement values of each run [24]. Unless otherwise noted, all subjects were required to have at least four minutes of unscrubbed frames [25].

Groups differed on the mean framewise displacement across scans before scrubbing and also on the number of unscrubbed frames (Table S1). To match groups on these two variables in our post-hoc analyses, we removed motion-prone patients (framewise displacement greater than 1.5 SD above the control mean; leaving 58 non-affective and 18 affective psychosis patients). For certain analyses, as a more austere measure, we also removed all subjects (patient or control) whose mean framewise displacement exceeded .08 mm so that the groups were again matched on this variable, similar to another prior study [26].

### Brain network partition

We used the Cole-Anticevic Brain Network partition, which divides parcels into 12 functional networks [11]. Functional networks were constructed from the 360 surface-based cortical parcels from the Glasser et al. atlas [27] plus an additional 358 volumetric subcortical parcels [11]. This partition includes four sensory networks: primary visual, secondary visual, somatomotor, and auditory (Fig. 1). There were 38 thalamic parcels, of which 22 were assigned to a sensory network (including 2 secondary visual, 2 somatomotor).

### Resting-state functional connectivity (RSFC) derivation

For each subject, we derived RSFC matrices via regularized partial correlation [28] and assessed each possible hyperparameter value via 10-fold cross-validation (hyperparameter range = 0-0.5 with increments of 0.001 from 0 to 0.01 and increments of .01 thereafter; see Supplementary Methods for details). Specifically, on each fold, a 718×718 regularized partial correlation matrix was formed from 90% of the time series. We then predicted the held-out portion of a time series of each parcel using this matrix along with the held-out time series of the remaining parcels (see Supplementary Methods). A hyperparameter value was considered optimal for a subject if it yielded a matrix that could most accurately predict the held-out time series across parcels and folds, where accuracy was assessed with the coefficient of determination (R^2^). An advantage to this method is that it yields FC estimates that are more accurate and more reliable than other multivariate approaches [13].

### Comparing groups on functional connectivity results

To determine each participant’s thalamo-cortical connectivity for a sensory network, we averaged all Fisher-z transformed connection weights between all 38 thalamic parcels and all cortical parcels of that network (yielding one value per subject). To determine each participant’s cortico-cortical connectivity value, we averaged Fisher-Z transformed connectivity weights between all cortical parcels of a network (again, yielding one value per subject). These averaged connectivity values were compared between groups using one-way ANOVAs, once for cortico-cortical and once for thalamo-cortical (Fig. 2). For pairwise comparisons of continuous quantities (including the cortico-cortical values, the thalamo-cortical values, and the proposed biomarker), we used Welch’s t-tests and Cohen’s d with Hedges’ correction (Hedges’ g) to account for sample size imbalances or potentially smaller sample sizes [29]. Statistical correction, when applied, was performed via Benjamini and Hochberg’s false discovery method (q<.05) [30]. Corrected p values were denoted with an “FDR” subscript.

**Fig. 2.**
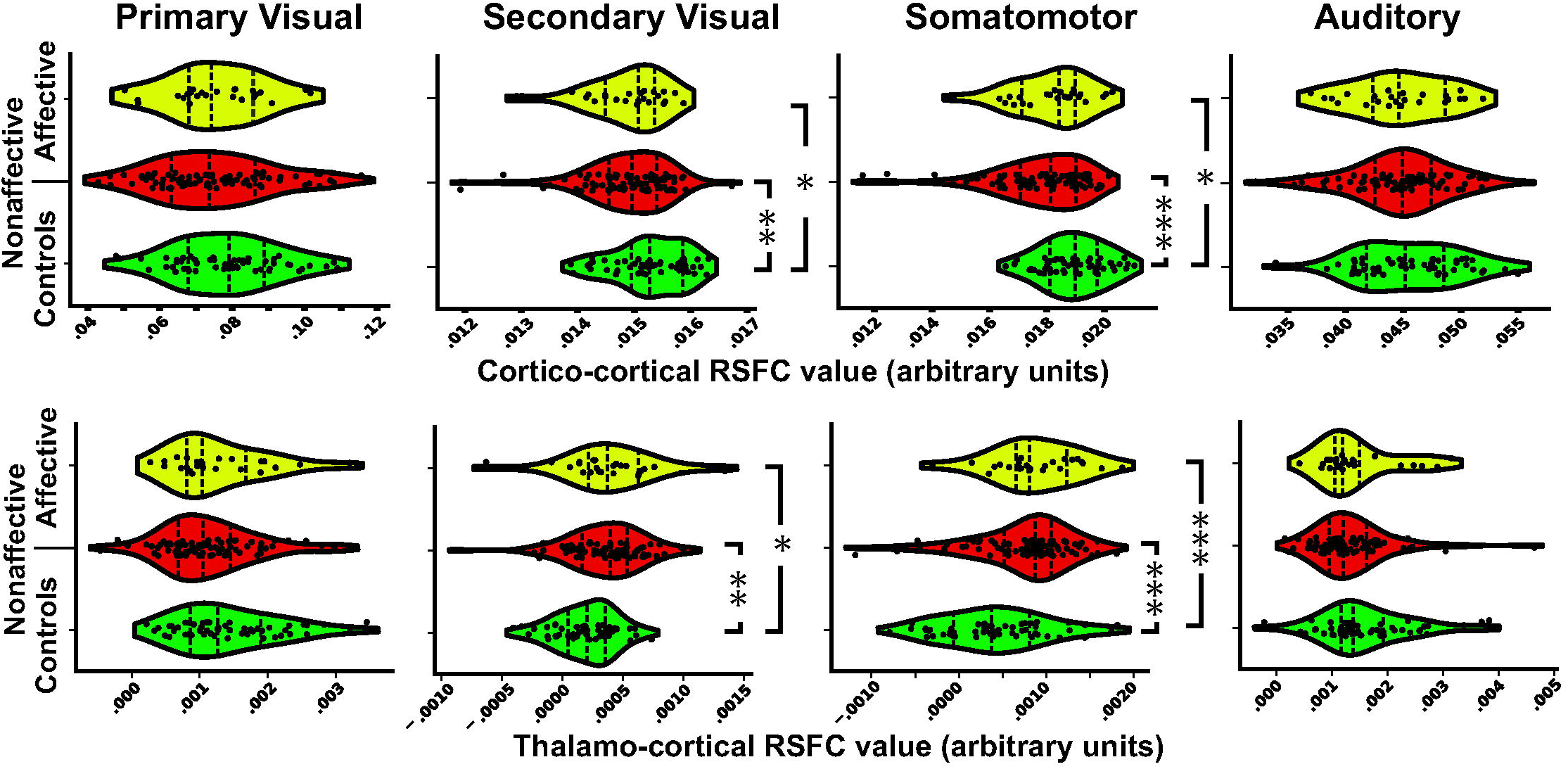
Group comparisons in cortico-cortical and thalamo-cortical connectivity for each sensory network. (Top) Cortico-cortical hypoconnectivity was found equally in non-affective and affective psychosis patients for both the somatomotor and visual2 networks. (Bottom) Thalamic hyperconnectivity was found equally in non-affective and affective psychosis patients for both the somatomotor and visual2 networks. Dotted lines indicate median and interquartile range of each distribution. Significant results are shown only for significant one-way ANOVAs and include FDR statistical correction. *p_FDR_<.05, **p_FDR_<.01, ***p_FDR_<.001

### Comparing groups using other data sets

We considered whether our proposed biomarker could be found in two independent data sets, each of which incorporated eyes-open resting-state data. The first was collected at Rutgers University and comprised 19 healthy controls and 22 chronically ill psychosis patients (14 schizophrenia, 1 schizoaffective disorder, 7 bipolar disorder); these data were collected on an older scanner (Tim Trio) with a different pulse sequence (e.g., MB 6, iPAT=2), a different scan duration (10 minutes; 765 TRs), and a different scrubbing threshold (0.3 mm) [31] The preprocessing has been described (ibid), with the only differences being that we also included subcortex (358 parcels), applied GSR, and derived the RSFC via graphical lasso, as above. The second independent data set was from the UCLA Consortium for Neuropsychiatric Phenomics–and compared people with ADHD (n=35) or schizophrenia (n=36), and healthy controls (n=93) [32]. The preprocessing steps for this single-band, legacy data set (with no T2-weighted structural image) have been described, but involved excluding GSR (using a variant of aCompCor instead), removing high motion scans and subjects, analyzing the whole brain in grayordinate space, and deriving the functional connectome via principal components multiple regression [for details see 33].

### Establishing the somato-visual marker via cross-validation and out-of-sample validation

To determine the predictive value of the somato-visual biomarker and to assign risk scores for each subject, we employed binary logistic regression and leave-one-site-out cross-validation (LOSOCV; four sites). Logistic regression was chosen because it is parsimonious, robust, and yields interpretable results [34, 35]. Sample size imbalances were minimized by using weighted logistic regression (so that sensitivity and specificity were given equal priority). LOSOCV was chosen because our goal was to determine if results could generalize to different populations and scanners, and since it has been used successfully in past studies [36, 37]. Note that, to prevent data leakage, the normalization terms (mean/SD) for the RSFC variable were derived from the training data only for each fold. We report key classification statistics from the LOSOCV, namely, sensitivity, specificity, positive predictive value, negative predictive value, balanced accuracy, and area under the ROC curve (AUC) (Table S2). AUC confidence intervals were provided via bootstrapping (1000 repetitions). Model performance was evaluated using all data with a 1-df Likelihood Ratio Test (LRT).

We also determined if the model built from the HCP data could predict the presence or absence of a psychotic disorder in the Rutgers and UCLA data sets described above. As before, the normalization terms were based on the training data only. We reported the same classification statistics as before (Table S3), and compared patients and controls on the risk scores in each held-out data set by using a one-sided Mann-Whitney U test.

### Determining the predictive value of RSFC by comparing it with neurocognition

We also examined whether the RSFC variable could improve upon neurocognition for classifying patients and controls. We utilized the “Q3A Memory” version of an auditory continuous performance task (ACPT) [38], in which participants heard two blocks of 90 pre-recorded letter sequences and were asked to indicate whenever they heard a “Q” followed by “A” four letters later. In a longitudinal study of clinical high risk patients, the hit rate from this task yielded one of the largest group differences (Cohen’s d=.7) between healthy controls and patients who went on to develop a psychotic disorder (n=264, n=89, respectively) [38]. Therefore, it was expected to provide a valid benchmark comparison. To consider whether the RSFC variable could improve upon this variable, we ran the weighted binary logistic regression on all subjects–once with the neurocognition variable by itself and once again with both variables included. We used a 1-df likelihood ratio test (LRT) to determine if the model improved by adding the RSFC variable.

### Test-retest reliability

To consider test-retest reliability, we: i) removed motion-prone patients as above; ii) computed the RSFC biomarker value separately for runs 1 and 2 (concatenated) and runs 3 and 4 (concatenated); and iii) calculated risk scores (across all subjects) at each time point by using weighted binary logistic regression. Finally, we probed whether the risk scores were correlated across the two time points and whether the RSFC biomarker was consistent across time points by using intraclass correlation (ICC(2,1); Supplementary Methods). Note that this ICC variant treats time point as a random factor to better generalize to longer retest intervals.

## Results

### Dysconnectivity of the somatomotor and secondary visual networks

With respect to cortico-cortical connectivity, the groups differed on the visual2 network (*F*(2,156)=5.6, *p*=0.004, η*²*=0.07; Fig. 2). Follow-up tests showed reduced connectivity in nonaffective patients relative to controls (t(123.6)=3.3, p*_FDR_*=0.004, g=0.54) and in affective patients relative to controls (*t*(42.2)=2.3, *p_FDR_*=0.04, *g*=0.57) but not between the two patient groups (*p*=.92, *g*=.02). There was also a group difference on cortico-cortical connectivity in the somatomotor network (*F*(2,156)=10.2, *p*=0.0001, η*²*=0.12). Follow-up tests showed reduced connectivity in each patient group relative to controls (non-affective: *t*(132.8)=4.8, *p_FDR_*<.0001, *g*=0.78; affective: *t*(37.7)=2.4, *p_FDR_*=0.03, *g*=0.63), but not between the two patient groups (*p*=.27, *g*=0.24). Groups did not differ on cortico-cortical connectivity of the primary visual network (*p*=.43, η*²*=.01) or auditory network (*p*=.52, η*²*=.01).

With respect to thalamo-cortical connectivity, the groups differed on the visual2 network (*F*(2,156)=6.2, *p*=0.003, η*²*=.07). Follow-up tests showed increased connectivity in nonaffective patients relative to controls (t(120.7)=3.4, p*_FDR_*=.0025, g=0.58) and in affective patients relative to controls (*t*(40.6)=2.5, *p_FDR_*=.023, *g*=0.64) but not between the two patient groups (*p*=.85). Groups also differed on thalamo-cortical connectivity in the somatomotor network (*F*(2,156)=13.3, *p*=5e-06, η*²*=0.15), with thalamic hyperconnectivity in nonaffective patients and affective patients relative to controls (*t*(131.5)=5.1, *p_FDR_*<.0001, *g*=0.83); t(37.5)=4.1, *p_FDR_*=.0003, *g*=1.07) but no patient group differences (*p*=.66, *g*=.10). There was some suggestion of patient thalamo-cortical *hypo*connectivity in the primary visual network (*F*(2,156)=3.3, *p*=.04, η*²*=0.04) and auditory network (*F*(2,156)=2.7, *p*=.07, η*²*=0.03), however, follow-up t-tests would not survive FDR correction. A split-half validation approach–which involved running these analyses on two equally split samples of controls and non-affective patients–yielded similar results (Supplementary Methods/Results), demonstrating robustness. When patient groups were combined, the prominent role of the somatomotor and visual2 networks in sensory dysconnectivity became even clearer (Fig. 3).

**Fig. 3.**
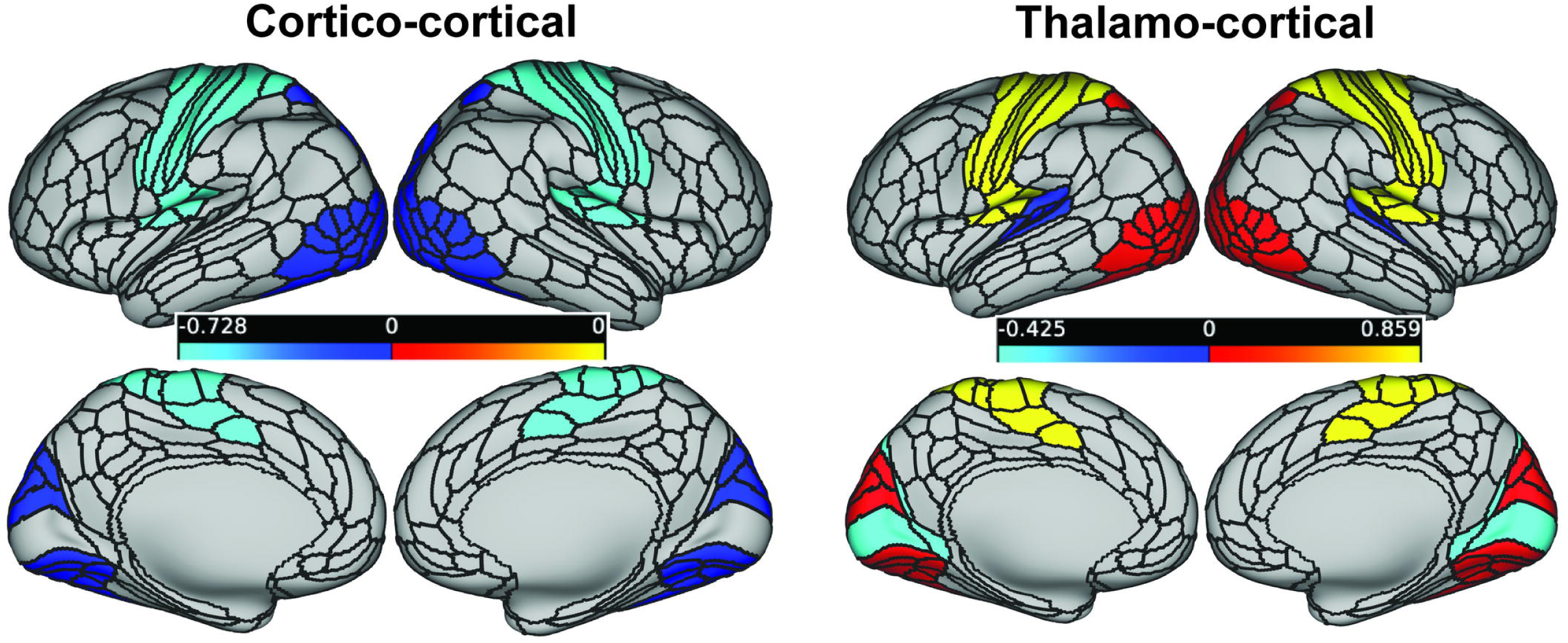
Network-wise group differences in connectivity across the four sensory networks using the combined patient sample (threshold FDR q<.05). Hedges’ g is shown in the legend. (Left) Across patients, cortico-cortical hypoconnectivity (in blue) was found in the visual2 and somatomotor networks but not in the other sensory networks. (Right) Across patients, thalamo-cortical hyperconnectivity (in yellow/red) was found with medium-large effect sizes in the visual2 and somatomotor network and hypoconnectivity was found with small effect sizes in the auditory and primary visual networks. Note that the two patient groups were combined because they did not differ for any network before correction for multiple comparisons.

### In-scanner motion cannot explain sensory dysconnectivity results

Patients in our sample exhibited more framewise displacement and had fewer usable frames after scrubbing (Table S1). This is a concern since increased motion can spuriously increase the observed connectivity between nearby regions and decrease the observed connectivity between distant regions; it can also degrade the MRI image quality [23, 39–41]. To better consider the effect of motion, we first examined whether motion correlated with each of the two variables (cortico-cortical connectivity, thalamo-cortical connectivity) for each of the two significant networks for each of the three groups. Across these 12 correlations, there was no significant effects after FDR correction (all |*r*|<.30), except for a negative correlation between framewise displacement and cortico-cortical visual2 connectivity in patients (*r*=-.32, *p_FDR_*=.04). To further consider the role of motion, we excluded all patients whose mean framewise displacement value before scrubbing was greater than 1.5 SD above the control mean (resulting in 58 non-affective, and 18 affective patients), so that groups were almost exactly matched on this variable and also on the number of unscrubbed frames (mean framewise displacement =.08 mm in each group; mean number of frames per group =1522-1548; both p>.3, both η*²* <.02). The four previously-significant one-way ANOVAs (two for visual2 and two for somatomotor) continued being significant (all η*²*>.05; all *p* <.03), with the control and nonaffective group showing the same differences as before (all *p*<.02; all *g*>.47).

### Medication cannot explain sensory dysconnectivity results

To consider the influence of medication, we first combined affective and non-affective patients since they did not differ in any of the connectivity measures described above. We then compared controls to the 22 psychosis patients (12 non-affective) who were naive to antipsychotics. The never-medicated patients had increased thalamo-cortical connectivity with the somatomotor and visual2 networks (*t*(32.6)=2.8, *p*=9e-03, *g*=0.76; *t*(51.0)=3.5, *p*=1e-03, *g*=0.78) and decreased cortico-cortical connectivity within the somatomotor and visual2 networks, although the last effect was only marginally significant (*t*(31.8)=3.2, *p*=.003, *g*=.91; *t*(47.4)=1.7, *p*<.10, *g*=0.39). We also directly compared patient groups with and without medication on these same connectivity measures. No differences emerged (all p>.17). Finally, to more fully consider antipsychotic effects, we probed for correlations between medication dose and connectivity values (cortico-cortical, thalamo-cortical) for these two networks. None of the four correlations reached significance (all |*r*|<.14, all *p*>.18, before correction). Thus, neuroleptics cannot explain the results.

### Combining across networks reveals larger group differences and reveals a new somato-visual biomarker for psychosis

Given the similar results for the visual2 and somatomotor networks, we averaged the two together to reduce noise and to potentially provide a stronger, overarching marker for psychosis. We combined patient groups, as above, and found larger effects than before (cortico-cortical: *t*(129.7)=5.0, *p*=1.3e-06, *g*=0.79; thalamo-cortical: *t*(141.3)=6.0, *p*=1.5e-08, *g*=0.90). Capitalizing on the fact that these two connectivity differences were approximately equal and opposite, we strove to generate an even stronger psychosis biomarker by normalizing these two values across all subjects, and subtracting the second from the first (thalamo-cortical - cortico-cortical). We found that the resulting “somato-visual” marker was elevated in patients compared to controls (*t*(136.2)=6.9, *p*=2e-10, *g*=1.05). A similar result would also arise if we were to use controls and only medication-naive patients (Fig. 4B). If we were to exclude motion-prone patients (58 non-affective and 18 affective in the combined sample), the effect strengthened (*t*(120.7)=7.1, *p*=9e-11, *g*=1.24). If we were to use an even more stringent threshold for all subjects (all having a mean framewise displacement <=.08 mm; 35 patients and 27 controls; similar to some prior studies [26]), the result was again strong (*t*(59.9)=4.2, *p*=1e-05, *g*=1.19).

**Fig. 4.**
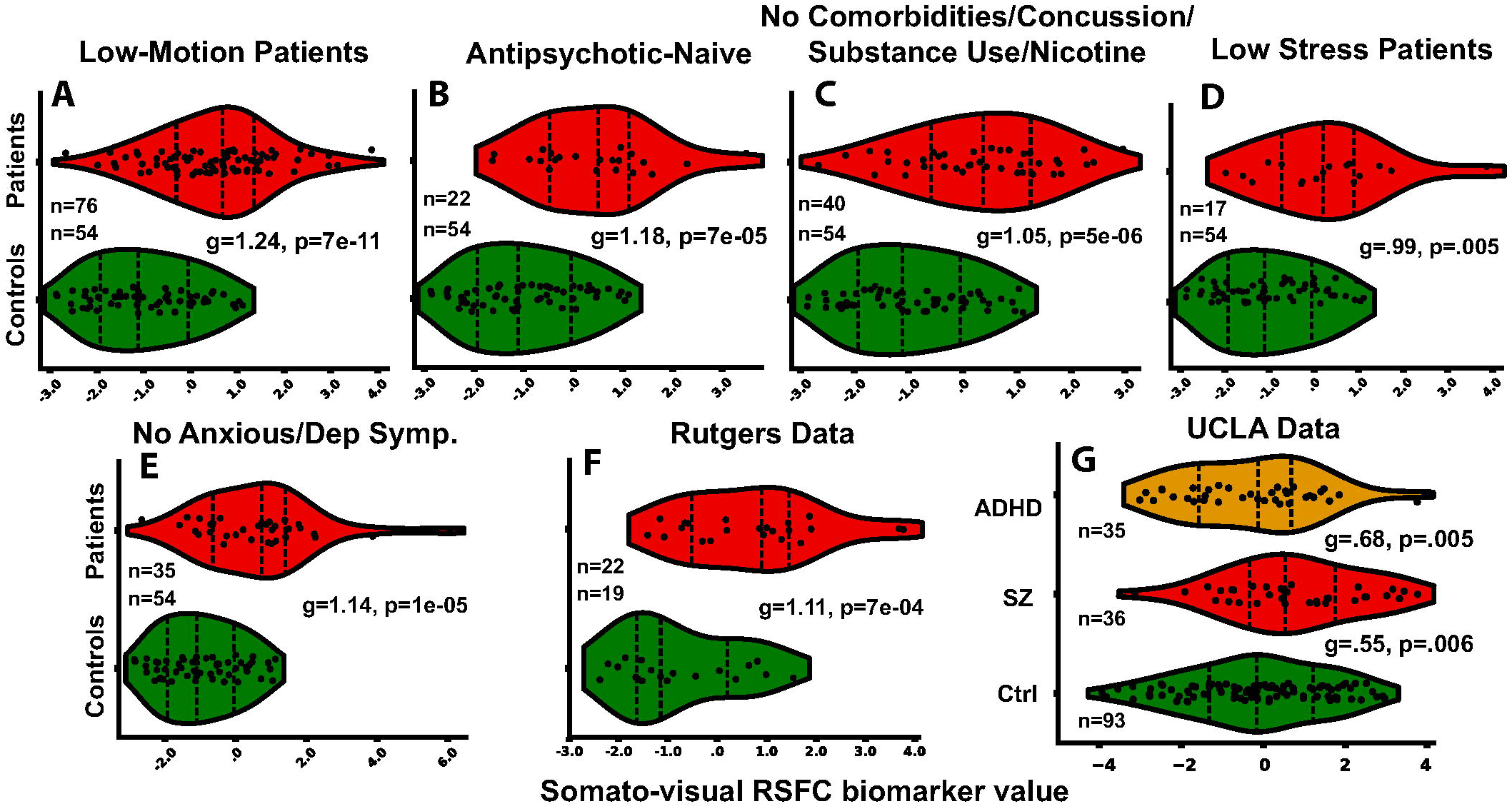
Demonstrating robustness of the somato-visual RSFC biomarker. (A-E) The biomarker could emerge when patients were evenly matched to controls on in-scanner motion, when they were medication-naive, when they had no comorbid conditions, concussive history, or substance/nicotine use, when they had no detectable levels of depression/anxiety in the two weeks prior to the scan, and when they had stress levels that were at or below the mean of the control group. (F, G) Groups differed on the biomarker in two additional data sets. Graphs A-F depict data with GSR and graphical lasso applied, except for the UCLA data set, which used aCompCor and principal components regression (see Methods).

### The somato-visual biomarker is robust to preprocessing strategy

To be credible, neuroimaging results should be robust to differences in preprocessing strategy [8]. This is important because patient/control RSFC differences have been shown to depend on preprocessing [25]. To this end, we re-ran the analyses with global signal regression (GSR; see Methods). The central results were qualitatively the same as before for the somato-visual biomarker (*t*(137.7)=6.9, *p*=1e-10, *g*=1.05; Supplementary Results; Fig. S2). Similar results also arose for a third preprocessing strategy, aCompCor, which uses the first five principal components from the white matter and ventricles (*t*(135.4)=6.8, *p*=3e-10, *g*=1.04) [20, 22]. Hereafter, we apply GSR since it did not qualitatively alter our findings and since others have argued that it offers the best strategy for denoising [20, 25, 36] and for revealing brain-behavior relationships [42].

### The somato-visual biomarker can be found with short scan durations

Although all subjects in our data sets were instructed to keep their eyes open, patients may more easily become drowsy in the scanner (e.g., due to sleep disturbances or medication), which can lead to RSFC nonstationarity [43] and create possible confounds in group comparisons, as noted by others [2]. Patients may also become more anxious in the bore, leading to drop-out bias for longer scan sessions. To consider whether the proposed biomarker can be recovered in a shorter duration, we re-ran the above analyses using only the first 5.5 minute scan and easily detected the biomarker (*t*(104.6)=5.9, *p*=5e-08, *g*=1.04; see Fig. S3). If we were to run only the very last run, the results would be weaker but still highly significant (*t*(127.7)=4.0, *p*=1e-04, *g*=0.66). Note that initial scans may be more accurate since sleepiness can worsen data quality and reliability, as demonstrated by others [33]. In either case, the data suggest that group differences can clearly emerge with a single run but may be more variable across runs (see also the test-retest reliability results below).

### The somato-visual biomarker cannot be explained by common confounds

Comorbidities and substance use pose a common confound in psychosis studies. Restricting our sample to patients who did not have a comorbid anxiety/mood/substance disorder, past mild concussion, or current nicotine use (n=40), we found that the biomarker value was higher in patients (*t*(74.2)=4.9, *p*=5e-06, *g*=1.05; Fig. 4C). Higher stress levels among patients may also be driving the results. However, if we include patients (n=17) with a stress level at or below the mean stress level of the controls using the Perceived Stress Scale raw scores [44], the biomarker value was again detectable (*t*(22.4)=3.1, *p*=5e-03, *g*=0.99; Fig. 4D). Heightened symptoms of depression/anxiety near the time of the scan might also explain why psychosis patients differ from healthy controls–a point made by others [45]. To rule out this possibility, we considered only patients who scored at the lowest possible level on all three items of this PANSS factor. The RSFC biomarker was once again elevated (*t*(54.2)=4.9, *p*=1e-05, *g*=1.14; Fig. 4E). The somato-visual biomarker did not differ between males and females in either patients or controls (both *p*>.21) and there was no correlation with IQ or parental educational attainment in either group (both *p*>.25, both |*r*|<.12). A patient’s race (black/white) can occasionally impact neuroimaging models of psychopathology [46] but we found no biomarker differences between black/white patients (p=.58; see Table S1) (Note that there were two few black controls (n=4) to meaningfully test this assertion in that group.) Finally, the magnitude of the patient/control differences did not depend on testing site, as determined by a 2 (group) x 4 (site) ANOVA (interaction: *F*(3,151)=1.2, *p*=.33).

### The somato-visual biomarker can be found in other data sets and with a clinical control group

If the aforementioned RSFC biomarker is robust and specific to psychosis, then it should be recoverable in different data sets and relative to a clinical control group. Using GSR and normalizing relative to that control sample, we found that patients in the Rutgers sample (22 psychosis, 19 healthy controls) again could be differentiated (*t*(39.0)=3.7, *p*=7e-04, *g*=1.11; Fig. 4F). To consider whether these results were a result of general psychopathology, we analyzed a third data set–the UCLA data set, which also contained ADHD participants (see Methods). The RSFC biomarker once again generated effects in the expected direction: Schizophrenia subjects (SZ) had higher values than healthy controls and ADHD patients (SZ vs Ctrl: *t*(65.6)=2.9, *p*=.006, *g*=.55; SZ vs. ADHD: *t*(69.0)=2.9, *p*=.005, *g*=.68; Fig. 4G). ADHD patients did not differ from controls (*t*(64.2)=0.5, *p*=.59, *g*=0.10).

### Four thalamic parcels undergird somato-visual functional dysconnectivity

If our results are specific to the somatomotor and visual2 networks, then we should be able to replicate the aforementioned findings using the four thalamic parcels of just those networks. We found such a result (*t*(122.8)=5.7, *p*=8e-08, *g*=0.91). If we were to require that patients exhibit less than .08 mm of average framewise displacement (to better isolate the signal from these four regions, the biomarker became even more apparent (four parcels: *t*(56.1)=4.7, *p*=2e-05, *g*=1.20). Thus even though we excluded 34 (89%) of the thalamic parcels, the biomarker could still be identified, though longer resting-state sessions or more reliable methods may be needed to establish the important role of these four parcels more definitively [47].

### The somato-visual biomarker can predict diagnostic status across sites and studies

Does the somato-visual biomarker have predictive value? Upon removing motion-prone patients (as above) and applying weighted binary logistic regression and leave-one-site-out cross-validation, we found that the RSFC biomarker could predict diagnostic status (sensitivity=.74, specificity=.70, AUC= 0.79, 95% CI=.76-81; *LRT*=43.2, *df*=1, *p*= 5e-11; Fig. 5A; see Table S2 for classification statistics). The results were nearly the same when motion-prone patients were included (Table S2).

**Fig. 5.**
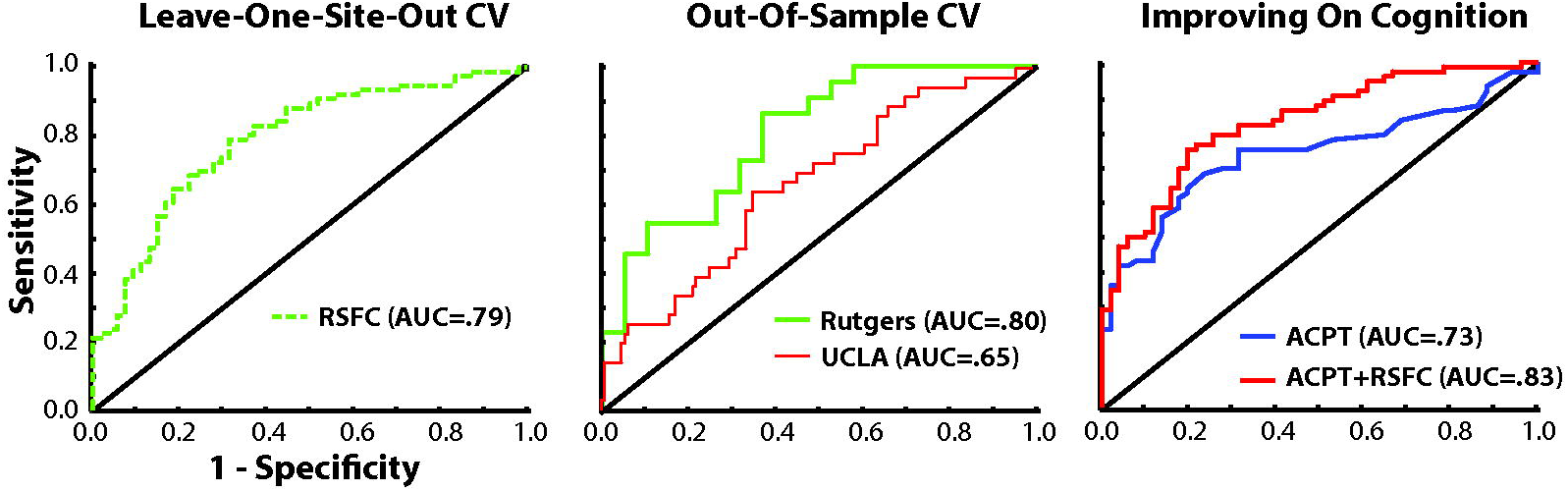
ROC curves. (A) The RSFC biomarker could distinguish 76 psychosis patients from 54 healthy controls using leave-one-site out cross-validation (LOSOCV). (B) A model constructed from the somato-visual biomarker variable in the HCP data could predict whether a participant had a psychotic disorder in two independent data sets (Rutgers: 19 controls, 22 psychosis patients; UCLA: 36 patients, 128 controls, including ADHD patients). (C) Using LOSOCV on the HCP data, the RSFC biomarker could boost group discrimination when added to the ACPT task (51 controls, 71 patients).

As an additional test, we considered whether a model constructed from the HCP data could classify subjects in the Rutgers and UCLA data sets (Fig. 5B; see Table S3 for classification statistics). We found that the model could indeed predict membership (psychosis or not) in each case (Rutgers: sensitivity=.68, specificity=.68, AUC=.80; Mann-Whitney test, U=3.3, p=5e-04; UCLA: sensitivity=.72, specificity=.47, AUC=.65, Mann-Whitney test, U=2.8, p=.002). The latter results were about the same if the ADHD controls were excluded (Table S3).

### The somato-visual biomarker can improve upon a well-established neurocognitive predictor

We next compared the predictive ability of the RSFC variable to the auditory CPT variable (ACPT-Q3A hit rate; Table S1; Fig. 5C). Using the same leave-one-site-out procedure as above (with motion-prone patients excluded), the CPT task by itself could discriminate the samples (sensitivity=.63, specificity=.80, *AUC*= .73, 95% CI=70-77). With the full sample, the results were significant when compared against the intercept model (*LRT*=44.8, *df*=1, *p*=2e-11). Importantly, when the fMRI measure was added (three regressors total, including intercept), the discrimination accuracy numerically improved (sensitivity=.76, specificity=.76, *AUC*= .83, 95% CI=77-86). Direct model comparison using the full sample confirmed that the RSFC measure improved group classification upon being added to the auditory CPT variable (*LRT* = 25.5, *df*=1, *p*=4e-07).

### Eleven minutes of resting-state can generate a moderately reliable somato-visual biomarker

We next considered whether the first pair of resting-state scans generated results resembling the second pair (test/retest interval =34.7 minutes). Incorporating only low-motion patients (using the same threshold as above; 54 controls, 74 patients), logistic regression risk scores were correlated across time points (*r*=.62, p=5e-14), and the biomarker had “moderate” intraclass correlation (*ICC*=.62, 95% CI=.49-.72, *p*=3e-14) using published benchmarks [48]. Supplementary results show that the ICC was hampered by the less-reliable thalamo-cortical connectivity values, especially at shorter scan durations.

## Discussion

We found that early psychosis patients exhibited thalamo-cortical hyperconnectivity and cortico-cortical hypoconnectivity with the visual2 and the somatomotor networks, but not with the auditory or primary visual networks. These results arose to a similar degree in affective and non-affective psychosis and did not depend on antipsychotic medication or in-scanner motion. Moreover, patient hypo- and hyperconnectivity patterns could be combined into a single overarching “somato-visual” biomarker. The marker could emerge within a single 5.5 minute scan, could not be explained by various common confounders, and could be revealed with two other data sets, one of which involved an ADHD control group. The marker could predict group membership across sites and studies, and could improve upon an auditory CPT task.

### Reassessing the granularity of “sensory dysconnectivity”

An implication is that it is painting with too broad a brush to say that there is “sensory dysconnectivity” or “visual dysconnectivity” in psychosis. The somatomotor and the visual2 networks were the primary driving factors; the auditory and primary visual networks issued forth much smaller group differences in cortico-cortical connectivity and potentially opposite group differences in thalamic connectivity (hypoconnectivity; see Fig. 3). Treating all visual regions or all sensory regions as the same will lead to an underestimation of the role of vision and sensation, respectively.

At the same time, our results suggest that sensory dysconnectivity may be minimally apparent at any one connection but may be clearly observed at the level of the network, when subtle local differences can be averaged over larger swaths of cortex [12]. This conclusion may explain why a recent machine-learning study of over 800 psychosis patients had difficulty in reliably discriminating psychosis patients and controls by using individual connection weights [49].

### What is the biological basis of a somato-visual biomarker?

Patients’ dysconnectivity patterns may be arising from NMDA glutamate receptor hypofunctioning [50, 51]. For example, in a double-blind, placebo controlled study, healthy participants who were administered ketamine–an NMDA receptor antagonist– exhibited thalamic hyperconnectivity with sensorimotor and higher-order visual cortex (e.g., postcentral gyrus, lingual gyrus) and this activity pattern more resembled early-stage psychosis patients than healthy controls [52]. The cortico-cortical functional hypoconnectivity of somatomotor and visual cortical regions has been observed in anti-NMDA receptor encephalitis, an autoimmune condition that produces symptoms resembling schizophrenia [53]. Structural connectivity differences may also play a role. Thalamo-cortical white matter tracts are more numerous for both somatosensory cortex (encompassed by the somatomotor network) and occipital cortex [54], presumably resulting from either novel white matter connections formed over the course of development or inadequate pruning during adolescence. Our results set the stage for more focused pathophysiological and pharmacological investigations.

### Somato-visual dysconnectivity as a biomarker for psychosis: Where do we go from here?

We already know how to diagnose psychosis, so why develop a biomarker based on current diagnosis? The biomarker’s simplicity, coupled with its large effect size, robustness, and generalizability, suggest that it may hold promise for predicting a future onset of psychosis among individuals at clinical high risk [55]. Adding neuroimaging to cognitive measures may offer an especially promising combination given how our biomarker could improve upon the ACPT (AUC=.73→.83). Moreover, while it remains unclear whether our biomarker is a cause or consequence of psychosis, it is at least conceivable that reversing hypo- or hyperconnectivity–either through neurostimulation, biofeedback, or pharmacological approaches–could reduce the likelihood or severity of illness [for a related example, see 56]. Finally, just as neuroimaging biomarkers of major depression have stratified patients into subgroups that respond to specific treatments [57], so too might the somato-visual biomarker in psychosis, although this will need to be tested on larger samples.

### Limitations and additional future directions

A limitation is that 11 minutes of resting-state yielded only a moderately reliable biomarker (ICC=.62). While this result is much better than what has been obtained with single edges from a FC matrix [mean ICC=.29, 58], future studies might consider using multi-echo fMRI or more advanced denoising strategies for image reconstruction [47, 59, 60]. Although the analyzed studies involved an eyes-open protocol, it is possible that patients more often fell asleep, which could explain their weaker cortico-cortical visual or somatomotor connectivity [61]. We consider this scenario unlikely since the strongest group differences occurred in the first resting-state scan, which began within the first few minutes of scanning. Nevertheless, future psychosis studies should more directly monitor wakefulness through measures such as eye-tracking. Another limitation is that we do not yet know the clinical or behavioral correlates of somato-visual dysconnectivity. We found no correlations with symptoms or functioning but the sample overall was extremely asymptomatic (see Table S1 and Supplementary results), necessitating future studies with more diverse patient types. In terms of behavioral performance, numerous visual abilities are abnormal in early-stage psychosis [62] including certain types of perceptual organization, which depend on the secondary visual network [63, 64]. Eye movements and visually guided reaching and grasping are also severely altered, and these alterations cannot easily be explained by antipsychotic medication [65–67]. A promising line of research would be to relate these behavioral impairments to connectivity patterns of the visual2 network [e.g., 64] or somatomotor network [e.g., 68, 69].

## Supporting information

Supplementary Methods/Results/Captions

## Acknowledgments

This work was supported by a K01MH108783 to BPK and a Hendershot pilot grant to BPK through the Psychiatry Department at the University of Rochester.

## Data and code availability

The HCP data are public (https://nda.nih.gov/edit_collection.html?id=2914). The UCLA data are located on OpenNeuro.org (accession number: ds000030), and so too are the Rutgers patient and control data (ds005073, ds003404, respectively). Preprocessing was done with fmriprep v.21.0.1 (www.fmriprep.org). Code for estimating RSFC and applying the Cole-Anticevic Brain Network Partition are on GitHub (https://github.com/ColeLab/ActflowToolbox/; https://github.com/ColeLab/ColeAnticevicNetPartition).

---

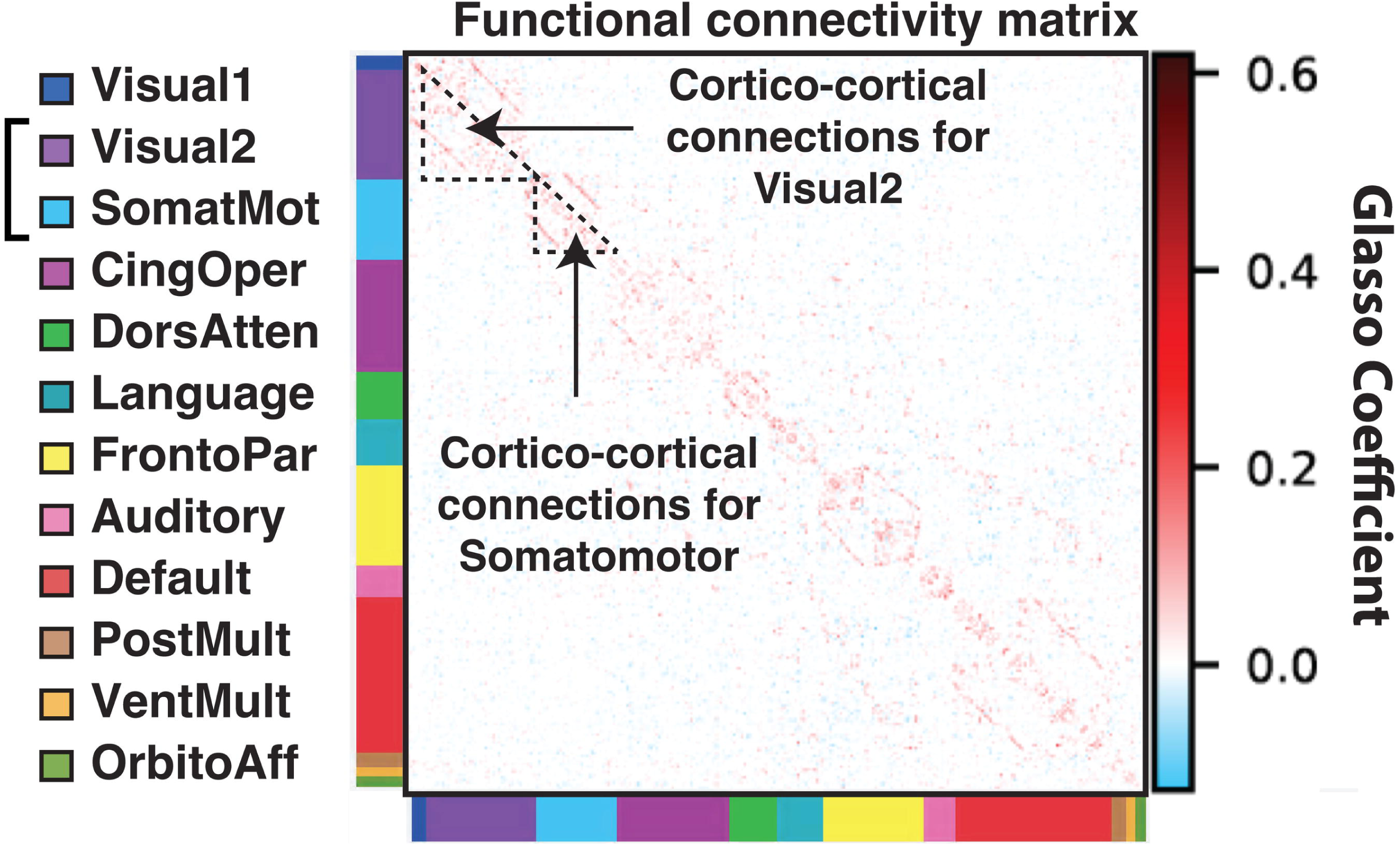

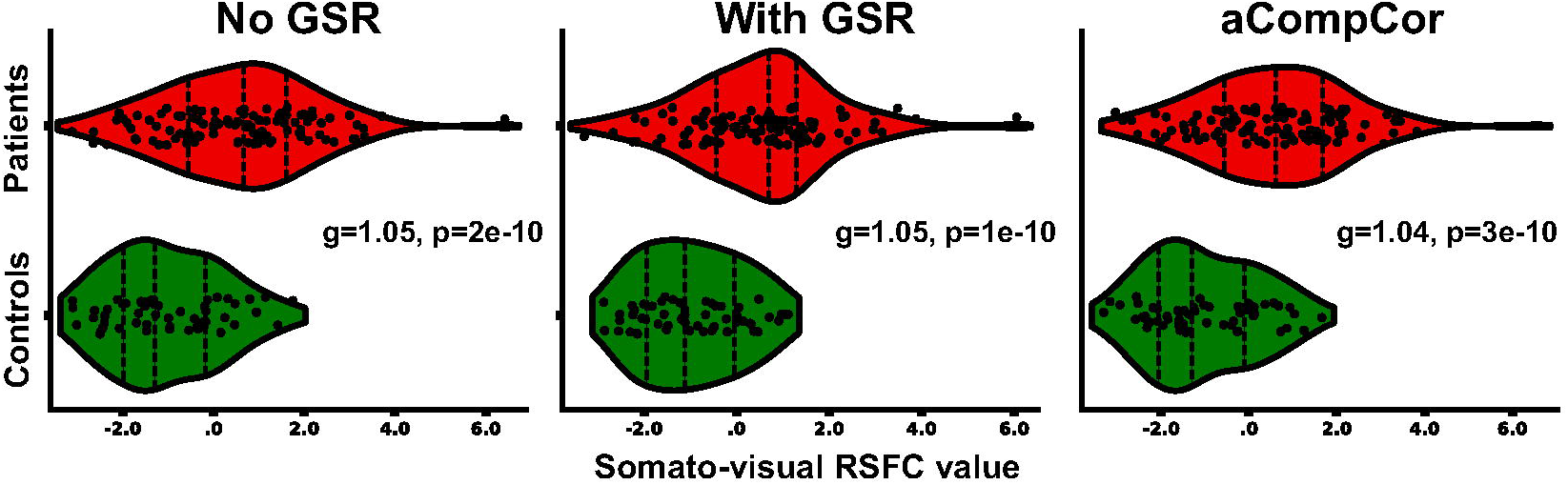

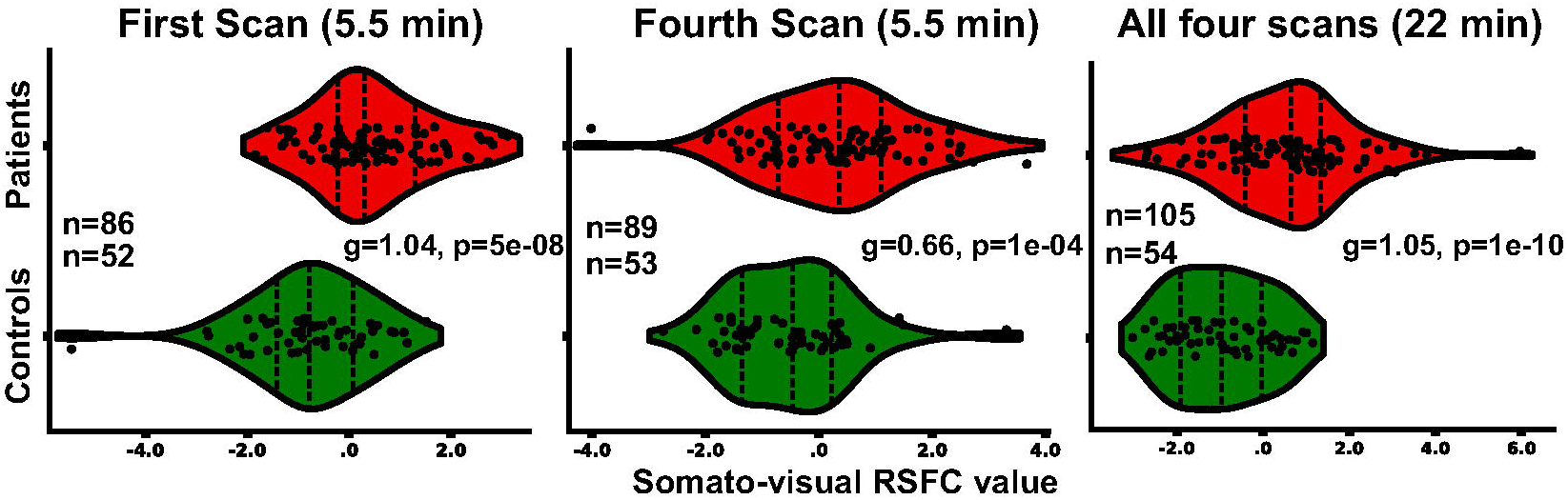

