## Supplementary Methods/Results/Captions for "Functional dysconnectivity of visual and somatomotor networks yields a simple and robust biomarker for psychosis"

**Supplementary Information**

**Supplementary Methods**

Details on the method are provided below but additional information regarding participant recruitment, inclusion/exclusion, clinical assessments, and study procedures can be found online (<https://nda.nih.gov/edit_collection.html?id=2914>).

*Participants*

The affective patients were diagnosed with major depression with psychosis or bipolar disorder with psychosis; the non-affective patients were diagnosed with schizophrenia, schizophreniform, schizoaffective, psychosis unspecified (formerly NOS), delusional disorder, or brief psychotic disorder. Healthy controls did not have a psychotic disorder or a first-degree family member with a schizophrenia spectrum disorder. Controls also did not have a recurrent major depressive disorder, an anxiety disorder that lasted for longer than 12 months or that required medication. Controls also did not have a history of psychiatric hospitalization and were not taking psychiatric medication at the time of testing. A mild or moderate substance use disorder was not disqualifying nor was having a past head injury so long as it did not impact brain functioning at the time of testing. Intellectual ability was gauged through the Full-Scale intelligence quotient (IQ) using the Wechsler Abbreviated Scale of Intelligence, Second Edition (WASI-II). The Hollingshead two-factor parental socioeconomic scale was used to measure parental educational attainment.

*MRI acquisition: HCP Early Psychosis*

##### The HCP-EP dataset was acquired on Siemens MAGNETOM Prisma 3T scanners. Subjects with usable neural data were as follows for each site: Beth Israel Deaconess Medical Center (21 patients, 8 controls), Indiana University (49 patients, 25 controls), Massachusetts General Hospital (7 patients, 9 controls), or McLean Hospital (28 patients, 12 controls). The functional scans were collected with the following parameters: time repetition (TR) = .8 s, echo time (TE) = 37 ms, flip angle = 52°, field of view (FOV) = 208 mm (oblique slice orientation), acceleration factor = 8. Subjects were asked to perform the resting-state scans with their eyes open (personal communication with PI M. Shenton on 3/15/24). T1w scans had the following parameters: TR = 2.4 s, TE = 2.22 ms, FOV = 256 mm, matrix = 256 × 256, sagittal plane, 0.8 mm slice thickness. T2w had the following parameters: TR = 3.2 s, TE = 563 ms, FOV = 256 mm, sagittal plane, 0.8 mm slice thickness.

*Minimal preprocessing (modified from fMRIPrep output)*

Auto-generated details of the minimal (initial) preprocessing steps are included below, with modifications for brevity and clarity. MRI minimal preprocessing was performed with fMRIPrep 21.0.1 (Esteban, [(Esteban et al., 2019; RRID:SCR_016216)](https://app.readcube.com/library/390abc1c-6162-479f-ad27-b1e1bbc3b86c/all?uuid=4391522685421485&item_ids=390abc1c-6162-479f-ad27-b1e1bbc3b86c:f48e1175-da87-4ed6-ac7b-365d6ced1594&options=%7B%22items%22%3A%7B%22390abc1c-6162-479f-ad27-b1e1bbc3b86c%3Af48e1175-da87-4ed6-ac7b-365d6ced1594%22%3A%7B%22suffix%22%3A%22%3B%20RRID%3ASCR_016216%22%7D%7D%7D), which is based on Nipype 1.6.1 [(Gorgolewski et al., 2011; RRID:SCR_002502)](https://app.readcube.com/library/390abc1c-6162-479f-ad27-b1e1bbc3b86c/all?uuid=06896064674389302&item_ids=390abc1c-6162-479f-ad27-b1e1bbc3b86c:72aa95f6-b05e-43dc-826b-696f0a6fe045&options=%7B%22items%22%3A%7B%22390abc1c-6162-479f-ad27-b1e1bbc3b86c%3A72aa95f6-b05e-43dc-826b-696f0a6fe045%22%3A%7B%22suffix%22%3A%22%3B%20RRID%3ASCR_002502%22%7D%7D%7D).

The T1-weighted (T1w) image was corrected for intensity non-uniformity (INU) with N4BiasFieldCorrection [(Tustison et al., 2010)](https://app.readcube.com/library/390abc1c-6162-479f-ad27-b1e1bbc3b86c/all?uuid=7667825608637988&item_ids=390abc1c-6162-479f-ad27-b1e1bbc3b86c:0b7a2454-7903-494e-b786-477194bdad96), distributed with ANTs 2.3.3 [(Avants et al., 2008, RRID:SCR_004757)](https://app.readcube.com/library/390abc1c-6162-479f-ad27-b1e1bbc3b86c/all?uuid=6133018638141536&item_ids=390abc1c-6162-479f-ad27-b1e1bbc3b86c:31b7393b-552d-4ea7-94af-2c7f2b48d6fe&options=%7B%22items%22%3A%7B%22390abc1c-6162-479f-ad27-b1e1bbc3b86c%3A31b7393b-552d-4ea7-94af-2c7f2b48d6fe%22%3A%7B%22suffix%22%3A%22%2C%20RRID%3ASCR_004757%22%7D%7D%7D), and used as T1w-reference throughout the workflow. The T1w-reference was then skull-stripped with a Nipype implementation of the antsBrainExtraction.sh workflow (from ANTs), using OASIS30ANTs as target template. Brain tissue segmentation of cerebrospinal fluid (CSF), white-matter (WM) and gray-matter (GM) was performed on the brain-extracted T1w using fast [(FSL 6.0.5.1:57b01774, RRID:SCR_002823, Zhang et al., 2001)](https://app.readcube.com/library/390abc1c-6162-479f-ad27-b1e1bbc3b86c/all?uuid=782401574679719&item_ids=390abc1c-6162-479f-ad27-b1e1bbc3b86c:1ca0f566-5f0f-42b4-8d78-9c8f84c4c57e&options=%7B%22items%22%3A%7B%22390abc1c-6162-479f-ad27-b1e1bbc3b86c%3A1ca0f566-5f0f-42b4-8d78-9c8f84c4c57e%22%3A%7B%22prefix%22%3A%22FSL%206.0.5.1%3A57b01774%2C%20RRID%3ASCR_002823%2C%20%22%7D%7D%7D). Brain surfaces were reconstructed using recon-all [(FreeSurfer 6.0.1, RRID:SCR_001847, Dale et al., 1999)](https://app.readcube.com/library/390abc1c-6162-479f-ad27-b1e1bbc3b86c/all?uuid=37856958192243206&item_ids=390abc1c-6162-479f-ad27-b1e1bbc3b86c:b82243f0-da61-428b-aa73-2ed1c4d3137a&options=%7B%22items%22%3A%7B%22390abc1c-6162-479f-ad27-b1e1bbc3b86c%3Ab82243f0-da61-428b-aa73-2ed1c4d3137a%22%3A%7B%22prefix%22%3A%22FreeSurfer%206.0.1%2C%20RRID%3ASCR_001847%2C%20%22%7D%7D%7D), and the brain mask estimated previously was refined with a custom variation of the method to reconcile ANTs-derived and FreeSurfer-derived segmentations of the cortical gray-matter of Mindboggle [(RRID:SCR_002438, Klein et al., 2017)](https://app.readcube.com/library/390abc1c-6162-479f-ad27-b1e1bbc3b86c/all?uuid=8605751722045167&item_ids=390abc1c-6162-479f-ad27-b1e1bbc3b86c:e10ebd05-59f6-4172-b2ff-4953ab626267&options=%7B%22items%22%3A%7B%22390abc1c-6162-479f-ad27-b1e1bbc3b86c%3Ae10ebd05-59f6-4172-b2ff-4953ab626267%22%3A%7B%22prefix%22%3A%22RRID%3ASCR_002438%2C%20%22%7D%7D%7D). Volume-based spatial normalization to two standard spaces (MNI152NLin2009cAsym, MNI152NLin6Asym) was performed through nonlinear registration with antsRegistration (ANTs 2.3.3), using brain-extracted versions of both T1w reference and the T1w template. The following templates were selected for spatial normalization: ICBM 152 Nonlinear Asymmetrical template version 2009c [Fonov et al. [(2009)](https://app.readcube.com/library/390abc1c-6162-479f-ad27-b1e1bbc3b86c/all?uuid=5944262540465014&item_ids=390abc1c-6162-479f-ad27-b1e1bbc3b86c:77bde660-2bb5-41db-9716-2ac67e34e79a&options=%7B%22items%22%3A%7B%22390abc1c-6162-479f-ad27-b1e1bbc3b86c%3A77bde660-2bb5-41db-9716-2ac67e34e79a%22%3A%7B%22suppressAuthor%22%3Atrue%7D%7D%7D), RRID:SCR_008796; TemplateFlow ID: MNI152NLin2009cAsym], FSL’s MNI ICBM 152 non-linear 6th Generation Asymmetric Average Brain Stereotaxic Registration Model [Evans et al. [(2012)](https://app.readcube.com/library/390abc1c-6162-479f-ad27-b1e1bbc3b86c/all?uuid=3595104653855369&item_ids=390abc1c-6162-479f-ad27-b1e1bbc3b86c:d5fe48e2-024a-43dd-93ee-544acb18927f&options=%7B%22items%22%3A%7B%22390abc1c-6162-479f-ad27-b1e1bbc3b86c%3Ad5fe48e2-024a-43dd-93ee-544acb18927f%22%3A%7B%22suppressAuthor%22%3Atrue%7D%7D%7D), RRID:SCR_002823; TemplateFlow ID: MNI152NLin6Asym].

For each of the 4 BOLD runs found per subject, the following preprocessing was performed. First, a reference volume and its skull-stripped version were generated by aligning and averaging 1 single-band reference (SBRef). Head-motion parameters with respect to the BOLD reference (transformation matrices, and six corresponding rotation and translation parameters) were estimated before any spatiotemporal filtering using mcflirt [(FSL 6.0.5.1:57b01774, Jenkinson et al., 2002)](https://app.readcube.com/library/390abc1c-6162-479f-ad27-b1e1bbc3b86c/all?uuid=7961808261674013&item_ids=390abc1c-6162-479f-ad27-b1e1bbc3b86c:3900fedf-7e51-4cf6-833a-048d10910c27&options=%7B%22items%22%3A%7B%22390abc1c-6162-479f-ad27-b1e1bbc3b86c%3A3900fedf-7e51-4cf6-833a-048d10910c27%22%3A%7B%22prefix%22%3A%22FSL%206.0.5.1%3A57b01774%2C%20%22%7D%7D%7D). The estimated fieldmap was then aligned with rigid-registration to the target EPI (echo-planar imaging) reference run. The field coefficients were mapped on to the reference EPI using the transform. BOLD runs were slice-time corrected to 0.348s (0.5 of slice acquisition range 0s-0.695s) using 3dTshift from AFNI [(Cox & Hyde, 1997, RRID:SCR_005927)](https://app.readcube.com/library/390abc1c-6162-479f-ad27-b1e1bbc3b86c/all?uuid=3872058228929053&item_ids=390abc1c-6162-479f-ad27-b1e1bbc3b86c:ac310819-ab11-47f9-ae42-9acfd0cad6f1&options=%7B%22items%22%3A%7B%22390abc1c-6162-479f-ad27-b1e1bbc3b86c%3Aac310819-ab11-47f9-ae42-9acfd0cad6f1%22%3A%7B%22suffix%22%3A%22%2C%20RRID%3ASCR_005927%22%7D%7D%7D). The BOLD reference was then co-registered to the T1w reference using bbregister (FreeSurfer) which implements boundary-based registration [(Greve & Fischl, 2009)](https://app.readcube.com/library/390abc1c-6162-479f-ad27-b1e1bbc3b86c/all?uuid=8670746334027437&item_ids=390abc1c-6162-479f-ad27-b1e1bbc3b86c:8bfd8a9a-8055-497d-a3fb-581d5e84f536). Co-registration was configured with six degrees of freedom. First, a reference volume and its skull-stripped version were generated using a custom methodology of fMRIPrep. Several confounding time-series were calculated based on the preprocessed BOLD, including framewise displacement (FD). FD was computed following Power (absolute sum of relative motions, Power et al. [(2014)](https://app.readcube.com/library/390abc1c-6162-479f-ad27-b1e1bbc3b86c/all?uuid=8563108027915829&item_ids=390abc1c-6162-479f-ad27-b1e1bbc3b86c:4aaa295f-0a5b-4a3c-b0c8-e3cb47a3a97d&options=%7B%22items%22%3A%7B%22390abc1c-6162-479f-ad27-b1e1bbc3b86c%3A4aaa295f-0a5b-4a3c-b0c8-e3cb47a3a97d%22%3A%7B%22suppressAuthor%22%3Atrue%7D%7D%7D)). The three global signals were extracted within the CSF, the WM, and the whole-brain masks. Additionally, a set of physiological regressors was extracted to allow for component-based noise correction [(CompCor, Behzadi et al., 2007)](https://app.readcube.com/library/390abc1c-6162-479f-ad27-b1e1bbc3b86c/all?uuid=9767810092557515&item_ids=390abc1c-6162-479f-ad27-b1e1bbc3b86c:1505f60d-96ef-48bf-9cdb-c65b0317ffc8&options=%7B%22items%22%3A%7B%22390abc1c-6162-479f-ad27-b1e1bbc3b86c%3A1505f60d-96ef-48bf-9cdb-c65b0317ffc8%22%3A%7B%22prefix%22%3A%22CompCor%2C%20%22%7D%7D%7D). Principal components were estimated after high-pass filtering the preprocessed BOLD time-series (using a discrete cosine filter with 128s cut-off) for the anatomical (aCompCor). For aCompCor, three probabilistic masks (CSF, WM and combined CSF+WM) were generated in anatomical space. The implementation differs from that of Behzadi et al. in that instead of eroding the masks by 2 pixels on BOLD space, the aCompCor masks are subtracted a mask of pixels that likely contain a volume fraction of GM. This mask was obtained by dilating a GM mask extracted from the FreeSurfer’s aseg segmentation, and it ensures components are not extracted from voxels containing a minimal fraction of GM. Finally, these masks were resampled into BOLD space and binarized by thresholding at 0.99 (as in the original implementation). Components are also calculated separately within the WM and CSF masks. For each CompCor decomposition, the k components with the largest singular values were retained. The confound time series derived from head motion estimates and global signals were expanded with the inclusion of temporal derivatives and quadratic terms for each . The BOLD time-series were resampled into standard space, generating a preprocessed BOLD run in MNI152NLin2009cAsym space. First, a reference volume and its skull-stripped version were generated using a custom methodology of fMRIPrep. The BOLD time-series were resampled onto the following surfaces (FreeSurfer reconstruction nomenclature): fsaverage. Grayordinates files [(Glasser et al., 2013)](https://app.readcube.com/library/390abc1c-6162-479f-ad27-b1e1bbc3b86c/all?uuid=2537550710172375&item_ids=390abc1c-6162-479f-ad27-b1e1bbc3b86c:3b80323d-a3e1-483a-abbb-5d7a80f3523e) containing 91k samples were also generated using the highest-resolution fsaverage as intermediate standardized surface space. The exact extension of the file used is ““_space-fsLR_den-91k_bold.dtseries.nii”. All resamplings can be performed with a single interpolation step by composing all the pertinent transformations (i.e. head-motion transform matrices, susceptibility distortion correction when available, and co-registrations to anatomical and output spaces). Gridded (volumetric) resamplings were performed using antsApplyTransforms (ANTs), configured with Lanczos interpolation to minimize the smoothing effects of other kernels [(Lanczos, 1964)](https://app.readcube.com/library/390abc1c-6162-479f-ad27-b1e1bbc3b86c/all?uuid=47632059458970233&item_ids=390abc1c-6162-479f-ad27-b1e1bbc3b86c:81dda848-f28e-449e-bcf3-9be5edd879cc). Non-gridded (surface) resamplings were performed using mri_vol2surf (FreeSurfer).

Many internal operations of fMRIPrep use Nilearn 0.8.1 [(Abraham et al., 2014, RRID:SCR_001362)](https://app.readcube.com/library/390abc1c-6162-479f-ad27-b1e1bbc3b86c/all?uuid=8133390566426703&item_ids=390abc1c-6162-479f-ad27-b1e1bbc3b86c:75c6a1c7-a149-4d7b-bdbf-65c24ccbe455&options=%7B%22items%22%3A%7B%22390abc1c-6162-479f-ad27-b1e1bbc3b86c%3A75c6a1c7-a149-4d7b-bdbf-65c24ccbe455%22%3A%7B%22suffix%22%3A%22%2C%20RRID%3ASCR_001362%22%7D%7D%7D), mostly within the functional processing workflow. For more details of the pipeline, see the section corresponding to workflows in fMRIPrep’s documentation.

*Additional preprocessing: Nuisance regression with anatomical CompCor*

To show robustness, we employed a third preprocessing strategy–dubbed aCompCor, which involves 10 physiological regressors formed from the first five principal components of the white matter and ventricle time series separately [(Ciric et al., 2017; Muschelli et al., 2014)](https://app.readcube.com/library/390abc1c-6162-479f-ad27-b1e1bbc3b86c/all?uuid=3819854363655697&item_ids=390abc1c-6162-479f-ad27-b1e1bbc3b86c:b5cbe656-d276-41c7-80c1-5662a4e7fa89,390abc1c-6162-479f-ad27-b1e1bbc3b86c:11363c98-28ac-4ac9-85f3-f0ac11190463) (see initial preprocessing steps described above). These 10 physiological regressors were used in addition to the same 24 motion regressors and 8 detrending/demeaning regressors mentioned in the main manuscript (for a total of 42 nuisance regressors).

*RSFC derivation: Additional details*

Each subject’s 718x718 RSFC matrix was generated by z-scoring the time series of each parcel and applying graphical lasso (“glasso”) as noted in the Methods. During the inversion of the covariance matrix, sparsity was imposed by applying an L1-penalty term to the model’s cost function. Note that the penalty term is proportional to the sum of the absolute values of the non-diagonal elements of the precision matrix and can be expressed as follows:

$\lambda_{1}\Sigma_{j\neq k}\left| \Theta_{jk} \right|$

where $\lambda_{1}$corresponds to the hyperparameter, and j and k correspond the row and column of the precision matrix $\Theta$. The precision matrix was then converted to a partial correlation matrix [(Peterson et al., 2023)](https://app.readcube.com/library/390abc1c-6162-479f-ad27-b1e1bbc3b86c/all?uuid=8602550053478801&item_ids=390abc1c-6162-479f-ad27-b1e1bbc3b86c:e2176b53-0788-4be9-9565-03f7e3b3fe3c):

$r_{AB|C}=-\Theta_{AB}\div\sqrt{\Theta_{AA}\Theta_{BB}}$

where r is the partial correlation coefficient for parcels A and B conditioned on C.

For each subject, we evaluated a range of possible hyperparameter values via 10-fold cross validation (λ_1_ hyperparameter range = 0-0.5 with increments of 0.001 from 0 to 0.005, then 0.01, and then increments of .01 thereafter). That is, for each fold, one-tenth of the time series was held-out and the remaining data were used to calculate an RSFC matrix using glasso. Each parcel’s held-out time series was then predicted in a linear regression model by using the connectivity values of all other parcels as beta coefficients and the held-out time series of the non-target parcels. For each fold, prediction accuracy for a held-out time series was assessed via the coefficient of determination (R^2^). A hyperparamater value was considered “optimal” for a subject if it yielded the largest coefficient of determination (R^2^) value on average across folds. An advantage to this method is that it yields FC estimates that are more accurate and more reliable than other multivariate approaches, when compared to either the structural connectome, the group-averaged functional connectome, or “ground-truth” connectome used in simulation analyses [(Peterson et al., 2023)](https://app.readcube.com/library/390abc1c-6162-479f-ad27-b1e1bbc3b86c/all?uuid=7608920923333454&item_ids=390abc1c-6162-479f-ad27-b1e1bbc3b86c:e2176b53-0788-4be9-9565-03f7e3b3fe3c).

After the optimal hyperparameter was chosen, we Fisher-Z transformed the partial correlation matrix to ensure that the results were approximately normally distributed. We then calculated thalamo-cortical and cortico-cortical connectivity values for each subject by averaging across connections as described in the Methods. An illustration of how we calculated connectivity values (in this case cortico-cortical) from the resulting connectome is shown in Fig. S1.

*“Split-half” validation results comparing cortico-cortical and thalamo-cortical connectivity values*

Split-half validation was used to evaluate the stability and robustness of cortico-cortical and thalamo-cortical connectivity when comparing groups [(Anderson & Magruder, 2017)](https://app.readcube.com/library/390abc1c-6162-479f-ad27-b1e1bbc3b86c/all?uuid=5427789171621941&item_ids=390abc1c-6162-479f-ad27-b1e1bbc3b86c:3e8505c0-54fc-489b-abf6-faccc34a33e2). To perform this procedure, we divided the non-affective and control sample in half, sorted each group in ascending order by their numeric ID, and assigned every other subject to the validation set and the remaining subjects to the discovery set. (Note that the affective psychosis sample was too small for split-half validation and split half validation was always done without GSR.) When a result was significant in the discovery data set, we re-ran the analysis in the validation data with a one-tailed test. A result that was significant on both tests was considered replicated and thus especially robust. Note that this procedure of “split half” validation is not to be confused with “out-of-sample cross-validation”, which involves building a logistic regression model on one sample and then testing it on another (see below).

*Test-retest reliability*

Following the convention of McGraw and Wong [(1996)](https://app.readcube.com/library/390abc1c-6162-479f-ad27-b1e1bbc3b86c/all?uuid=26687248076890624&item_ids=390abc1c-6162-479f-ad27-b1e1bbc3b86c:aee0a968-4ec5-4644-958c-c326e5b9a46a&options=%7B%22items%22%3A%7B%22390abc1c-6162-479f-ad27-b1e1bbc3b86c%3Aaee0a968-4ec5-4644-958c-c326e5b9a46a%22%3A%7B%22suppressAuthor%22%3Atrue%7D%7D%7D), we implemented a two-way random effects model (“ICC(2,1)”) that considered absolute agreement on a single measure, as also recommended by others [(see also, Koo & Li, 2016)](https://app.readcube.com/library/390abc1c-6162-479f-ad27-b1e1bbc3b86c/all?uuid=1865714233824194&item_ids=390abc1c-6162-479f-ad27-b1e1bbc3b86c:f2ca35df-401c-4966-a6ec-20473781c8ea&options=%7B%22items%22%3A%7B%22390abc1c-6162-479f-ad27-b1e1bbc3b86c%3Af2ca35df-401c-4966-a6ec-20473781c8ea%22%3A%7B%22prefix%22%3A%22see%20also%2C%20%22%7D%7D%7D). ICC is therefore defined as:

MS_R_ - MS_E_

___________________________

MS_R_ + (k-1)MS_E_ + (k/n)(MS_C_ - MS_E_)

##### In this notation, MS_R_= mean squares between subjects (variability of the RSFC biomarker across subjects); MS_E_ = mean square for error; MS_C_ = mean square for time points (variability of the biomarker across time points); n = number of subjects; and k = number of time points.

The end of the first two resting-state scans was separated by the end of the second pair of resting-state scans by an average of 35 minutes (SD=2.6 min), being intervened by field maps, diffusion scans, and T1w/T2w scans. The end of the first resting-state scan was separated from the end of the last resting-state scan by 47 minutes (SD=2.7 min).

**Supplementary Results**

*Dysconnectivity of the somatomotor and secondary visual networks could be replicated in split-half validation*

To show robustness, we considered thalamo-cortical and cortico-cortical connectivity using split-half validation (see above for Methods). With respect to cortico-cortical connectivity, we compared healthy controls and non-affective psychosis patients. In the secondary visual network, the non-affective psychosis patients exhibited cortico-cortical hypoconnectivity in the discovery sample (t(47.1)=3.5, p=0.001, g=0.90) and to a marginal extent in the validation sample (*t*(65.0)=1.3, *p*=0.09, *g*=0.31). In the somatomotor network, the non-affective psychosis patients again exhibited hypoconnectivity in the discovery sample (*t*(65.4)=2.8, *p*=0.007, *g*=0.63) and validation sample (*t*(65.0)=4.1, *p*=0.0001, *g*=0.94). There were no cortico-cortical connectivity differences in either the primary visual or auditory networks for either the discovery or validation samples (both *p*>.12).

With respect to thalamo-cortical connectivity, patients exhibited hyperconnectivity in the secondary visual network in the discovery (*t*(54.1)=2.1, *p*=0.04, *g*=0.52) and validation samples (*t*(64.0)=2.7, *p*=0.004, *g*=0.64). There was thalamic hyperconnectivity with the somatomotor network in the discovery (*t*(62.4)=4.7, *p*<.0001, *g*=1.10) and validation data (*t*(65.0)=2.6, *p*=0.005, *g*=0.61). Although there was hypoconnectivity in the visual1 and auditory networks (both *p*<.05, both *g*>.66), the effects did not remain significant or marginally significant in the validation.

*Results with different preprocessing strategies: GSR and aCompCor*

Results were similar for different preprocessing strategies. Using GSR, patients as a whole had decreased cortico-cortical connectivity with visual2 and somatomotor networks (visual2: *t*(102.8)=3.7, *p*=0.0003, *g*=0.55; somatomotor: *t*(124.8)=5.3, *p*=6e-07, *g*=0.69) and increased thalamo-cortical connectivity for the same networks (visual2: *t*(96.7)=2.4, *p*=0.016, *g*=0.39); somatomotor: *t*(118.0)=5.7, *p*=1e-07, *g*=0.82). In patients, there was small thalamo-cortical hypoconnectivity with the primary visual network (t(74.4)=2.2, p=0.03, g=0.39) and the auditory network (*t*(105.6)=2.1, *p*=0.04, *g*=0.36).

Using aCompCor, patients as a whole had decreased cortico-cortical connectivity with visual2 and somatomotor networks (visual2: *t*(99.2)=3.7, *p*=0.0007, *g*=0.56; somatomotor: *t*(133.0)=5.7, *p*=8e-08, *g*=0.73) and increased thalamo-cortical connectivity for the same networks (visual2: *t*(96.9)=-3.7, *p*=0.0003, *g*=0.58); somatomotor: *t*(119.2)=-6.0, *p*=2e-08, *g*=0.86). In patients, there was small thalamo-cortical hypoconnectivity with the primary visual network (*t*(73.7)=2.7, *p*=0.008, *g*=0.46) and the auditory network (*t*(95.3)=2.4, *p*=0.02, *g*=0.42).

The biomarker variable was strong with GSR (*t*(137.7)=6.9, *p*=1e-10, *g*=1.05), without GSR *t*(136.2)=6.9, *p*=2e-10, *g*=1.05), and with aCompCor (*t*(135.4)=6.8, *p*=3e-10, g=1.04) (see Fig. S2).

*Additional results with shorter scan durations*

As noted in the main manuscript, we re-ran our analyses using the first 5.5 minute resting state scan and achieved strong results (Fig. S3). A caveat is that many more subjects were removed since they did not have at least four minutes of unscrubbed frames when we examined only a single scan (2 controls, 18 non-affective, 4 affective). If we were to instead require only 2 minutes of unscrubbed frames (removing only 8 non-affective and 1 affective patients, the biomarker would weaken somewhat but still be strong (*t*(119.1)=5.9, *p*=4e-08, *g*=0.96). We also re-ran the fourth scan (removing 1 control, 16 non-affective, and 3 affective patients). The results were significant but weaker overall (*t*(127.7)=4.0, *p*=1e-04, *g*=0.66). If we required only 2 minutes of unscrubbed frames (removing 4 non-affective, 1 affective), similar results were obtained (*t*(136.1)=4.4, *p*=2e-05, *g*=0.68). Thus, the number of subjects removed is not obviously connected to the strength of the group differences. We present evidence further below that indicates that results with a single-scan are less reliable and more variable, as compared to when using two or four concatenated scans.

*Results with four thalamic parcels: Longer scans may be needed to show the effect*

As noted in the manuscript, four thalamic parcels discriminated groups when removing the remaining 34 thalamic parcels, especially if we strictly controlled for motion. A caveat is that this effect diminished if we were to use only one scan (*t*(117.8)=3.9, *p*=1e-04, *g*=0.67), including when motion-prone patients were removed (t(115.7)=3.6, *p*=5e-04, *g*=0.63). This suggests, not surprisingly, that longer scan durations are probably necessary to reliably monitor activity and identify a biomarker from specific thalamic parcels.

*Additional test-retest reliability results: reduced reliability of subcortex hampers ICC, especially for shorter scan durations*

To better understand why the ICC for the somato-visual marker was not higher, we separately examined the ICCs of the raw cortico-cortical and thalamo-cortical connectivity values (averaged across networks, as in the main manuscript). We found that the ICC was higher for the cortico-cortical connectivity values (*ICC*=.79, 95% CI= [.71, .85], *p*<3e-16) but lower for the thalamo-cortical connectivity values (*ICC*=.47 95% CI=[.32, .60], *p*<3e-16). This suggests that the ICC was of only moderate magnitude because of poor reliability of subcortex.

To consider the impact of scan duration, we conducted the same analyses using only the first and last resting-state scans (53 controls, 74 patients). The results became predictably worse: Risk scores were less correlated across time points (*r*=.49, *p*=4e-08), and the biomarker had poor intraclass correlation (*ICC*=.46 [.31, .60], *p*=1e-07). The ICC worsened for cortico-cortical connectivity (*ICC*=.63 [0.51, 0.73], *p*=2e-14) but worsened even more for thalamo-cortical connectivity (*ICC*=.21 [.03, .38], *p*=.01). This suggests that poor reliability of subcortex drags down the biomarker ICC, especially for shorter scan durations.

*Relationships with symptoms and functioning*

The somato-visual marker did not correlate with occupational or social functioning, as derived from the Global Assessment of Functioning scale (both *p*>.14, both |*r*|<.15, before correction). There were also no correlations with positive, negative, disorganized, depressive, or excitement symptoms (all |*r*|<.11, all *p*>.29, before correction). It should be emphasized, however, that this was a highly asymptomatic sample (Table S1): In the most relevant symptom categories (positive, negative, disorganized), fewer than 2% of patients had at least moderate symptoms (mean PANSS item>=4) and fewer than 14% had at least mild symptoms (mean PANSS item>=3). As shown in Table S1, the two patient groups were also highly functioning, with both falling in the 60s and 70s.

### **Supplementary Tables**

**Table S1. Demographic and clinical data for HCP sample.** Parental educational attainment was based on the Hollingshead Index and was averaged between parents. Framewise displacement was calculated before scrubbing. The number of unscrubbed frames was based on a 0.2 mm framewise displacement scrubbing threshold and was calculated after temporal filtering (see Methods). The RSFC biomarker (arbitrary units) was calculated with global signal regression (GSR). Total antipsychotic exposure was measured in months at the time of testing. Occupational and social functioning were measured with the Global Assessment of Functioning Scale (GAF) (APA, 2013). PANSS scores provide the group mean of the factor score averaged across items (minimum=1.0, maximum = 7.0), where 1 = symptom absent and 2 = symptom questionably present. The group comparison column shows results from a one-way ANOVA, except for categorical variables, which used a Chi-Square test. Follow-up pairwise tests (uncorrected) were conducted when three groups differed significantly on a one-way ANOVA or Chi-Square test. There was a small amount of missing data for handedness (1 non-affective), IQ (1 in each group), the ACPT-Q3A task (3 controls, 4 non-affective, 1 affective), social functioning (1 affective), and PANSS (1 affective).

<<Table S1 here>>

**Table S2**. **Classification statistics for leave-one-out cross-validation**. The left column shows classification results that emerged upon applying leave-one-out cross-validation with weighted binary logistic regression. We performed the analysis once on the APCT-Q3A task alone, once on the RSFC variable alone, and once using both variables combined. In the column showing “still” patients, motion-prone patients were removed from all three analyses so that the two groups were matched closely on in-scanner motion (76 patients, 54 controls). In the column showing all patients, motion-prone patients were not removed in this way (105 patients, 54 controls). Note that there was no task data for 5 patients and 3 controls (still patients: 71; all patients: 100) and so these individuals were excluded from all rows that involved the ACPT-Q3A variable.

<<Table S2 here >>

**Table S3**. **Classification statistics for out-of-sample validation with the Rutgers and UCLA data sets**. The first column of data shows classification results that emerged upon applying the model constructed from the HCP data to the Rutgers data set. The second and third column show classification results that emerged upon applying the model to the UCLA data set with ADHD patients included or with ADHD patients excluded. The Rutgers patient sample included schizophrenia (n=14), schizoaffective (n=1), and bipolar disorder patients with psychotic features (n=7). The UCLA data set included 36 schizophrenia patients, 35 ADHD patients and 93 healthy controls. Note that the UCLA data was a legacy data set (with no T2-weighted structural image) and did not have GSR or graphical lasso applied; thus, it provides a conservative estimate of the classifier’s utility.

<<Table S3 here >>

### **Supplementary Figures**

<<Fig. S1 here>>

**Fig. S1. Sample FC matrix with parcels sorted by functional (color-coded) network.** Cortico-cortical connectivity values were generated for each person and each sensory network by averaging the connection weights between cortical parcels of a network. Here, the cortical parcels pertaining to the somatomotor and secondary visual networks are outlined. Note that, in graphical lasso, L1 regularization shrinks less informative coefficients to exactly zero, resulting in a sparse matrix.

<<Fig. S2 here>>

**Fig. S2**. **The somato-visual biomarker values for three different preprocessing strategies.** There were 54 controls and 105 patients in each panel. The “No GSR” graph involved 40 regressors (including 8 detrend/demean regressors); the “GSR” graph involved 44 regressors (which also includes 4 global signal regressors), and the “aCompCor” involved 42 regressors (including 10 physiological regressors but no global signal regressors). As can be seen, the results were about the same for each type of preprocessing. We adopted GSR in the main manuscript simply because others have argued that it provides the best denoising strategy and because it was considered potentially more likely to recover correlations with symptoms.

<<Fig. S3 here>>

**Fig. S3. Somato-visual biomarker results for scan 1, scan 4, or all scans.** Note that the number of subjects varied from scan to scan because subjects needed to have at least four minutes of unscrubbed frames, and this value depended on the number of scans concatenated.

[Lanczos, C. (1*96*4). A Precision Approximation of the Gamma Function. *Journal of the Society for Industrial and Applied Mathematics Series B Numerical Analysis*, *1*(1), 86–96. https://doi.org/10.1137/0701008](https://app.readcube.com/library/?style=American%20Psychological%20Association%207th%20edition+%7B%22language%22:%22en-US%22%7D)

[McGraw, K. O., & Wong, S. P. (1996). Forming Inferences About Some Intraclass Correlation Coefficients. *Psychological Methods*, *1*(1), 30–46. https://doi.org/10.1037/1082-989x.1.1.30](https://app.readcube.com/library/?style=American%20Psychological%20Association%207th%20edition+%7B%22language%22:%22en-US%22%7D)

[Muschelli, J., Nebel, M. B., Caffo, B. S., Barber, A. D., Pekar, J. J., & Mostofsky, S. H. (2014). Reduction of motion-related artifacts in resting state fMRI using aCompCor. NeuroImage, 96, 22–35. https://doi.org/10.1016/j.neuroimage.2014.03.028](https://app.readcube.com/library/?style=American%20Psychological%20Association%207th%20edition+%7B%22language%22:%22en-US%22%7D)

[Peterson, K. L., Sanchez-Romero, R., Mill, R. D., & Cole, M. W. (2023). *Regularized partial correlation provides reliable functional connectivity estimates while correcting for widespread confounding*. https://doi.org/10.1101/2023.09.16.558065](https://app.readcube.com/library/?style=American%20Psychological%20Association%207th%20edition+%7B%22language%22:%22en-US%22%7D)

[Power, J. D., Mitra, A., Laumann, T. O., Snyder, A. Z., Schlaggar, B. L., & Petersen, S. E. (2014). Methods to detect, characterize, and remove motion artifact in resting state fMRI. NeuroImage, 84, 320–341. https://doi.org/10.1016/j.neuroimage.2013.08.048](https://app.readcube.com/library/?style=American%20Psychological%20Association%207th%20edition+%7B%22language%22:%22en-US%22%7D)

[Tustison, N. J., Avants, B. B., Cook, P. A., Zheng, Y., Egan, A., Yushkevich, P. A., & Gee, J. C. (2010). N4ITK: Improved N3 Bias Correction. *IEEE Transactions on Medical Imaging*, *29*(6), 1310–13*20*. https://doi.org/10.1109/tmi.2010.2046908](https://app.readcube.com/library/?style=American%20Psychological%20Association%207th%20edition+%7B%22language%22:%22en-US%22%7D)

[Zhang, Y., Brady, M., & Smith, S. (2001). Segmentation of Brain MR Images Through a Hidden Markov Random Field Model and the Expectation-Maximization Algorithm. IEEE Transactions on Medical Imaging, 20(1), 45. https://doi.org/10.1109/42.906424](https://app.readcube.com/library/?style=American%20Psychological%20Association%207th%20edition+%7B%22language%22:%22en-US%22%7D)
